# Quantifying heterogeneity in SARS-CoV-2 transmission during the lockdown in India

**DOI:** 10.1101/2020.09.10.20190017

**Authors:** Nimalan Arinaminpathy, Jishnu Das, Tyler H. McCormick, Partha Mukhopadhyay, Neelanjan Sircar

## Abstract

The novel SARS-CoV-2 virus shows marked heterogeneity in its transmission. Here, we used data collected from contact tracing during the lockdown in Punjab, a major state in India, to quantify this heterogeneity, and to examine implications for transmission dynamics. We found evidence of heterogeneity acting at multiple levels: in the number of potentially infectious contacts per index case, and in the per-contact risk of infection. Incorporating these findings in simple mathematical models of disease transmission reveals that these heterogeneities act in combination to strongly influence transmission dynamics. Standard approaches, such as representing heterogeneity through secondary case distributions, could be biased by neglecting these underlying interactions between heterogeneities. We discuss implications for policy, and for more efficient contact tracing in resource-constrained settings such as India. Our results highlight how contact tracing, an important public health measure, can also provide important insights into epidemic spread and control.

## Introduction

There is increasing recognition of pronounced heterogeneity in the transmission of SARS-CoV-2: that is, that the majority of transmission events appear to be caused only by a small proportion of infected individuals (*1-4*). Previous modelling work has highlighted the importance of heterogeneity in the emergence of novel pathogens (*5*), as well as its implications for herd immunity to SARS-CoV-2 (*3, 6*). Understanding heterogeneity can also have important implications for control, if interventions can be targeted at those most likely to contribute to transmission (*7*). The need to streamline resources in this way is especially pressing in low- and middle-income settings, given fears that healthcare services in these settings would be particularly challenged by SARS-CoV-2 (*8*). Here, we analysed data collected from contact tracing during the lockdown in Punjab, a major Indian state, to understand heterogeneity of transmission in this setting, and its implications for control.

## Epidemiological context

Punjab, a state in India of about 30 million inhabitants, went into lockdown from 1st April to May 26th (Fig.1A). As elsewhere in India, the lockdown heavily restricted the movement of populations, in most cases to their homes and immediate neighborhoods. Travelling outside the house required a special pass, except for essential activities which were also restricted to certain times of the day. The Government of Punjab conducted intensive contact tracing during this time, amongst all known contacts of positive cases, and regardless of symptom status. Due to the ease of tracking individuals during the lockdown, 95% of high-risk contacts (defined as those having face-to-face conversation for at least 15 minutes) could be effectively traced and tested. Overall, this data constitutes the census of all infected persons and their contacts in the state; owing to the lockdown conditions, it affords a unique opportunity to measure contacts with greater accuracy than would be possible during normal economic activity.

**Figure 1:**
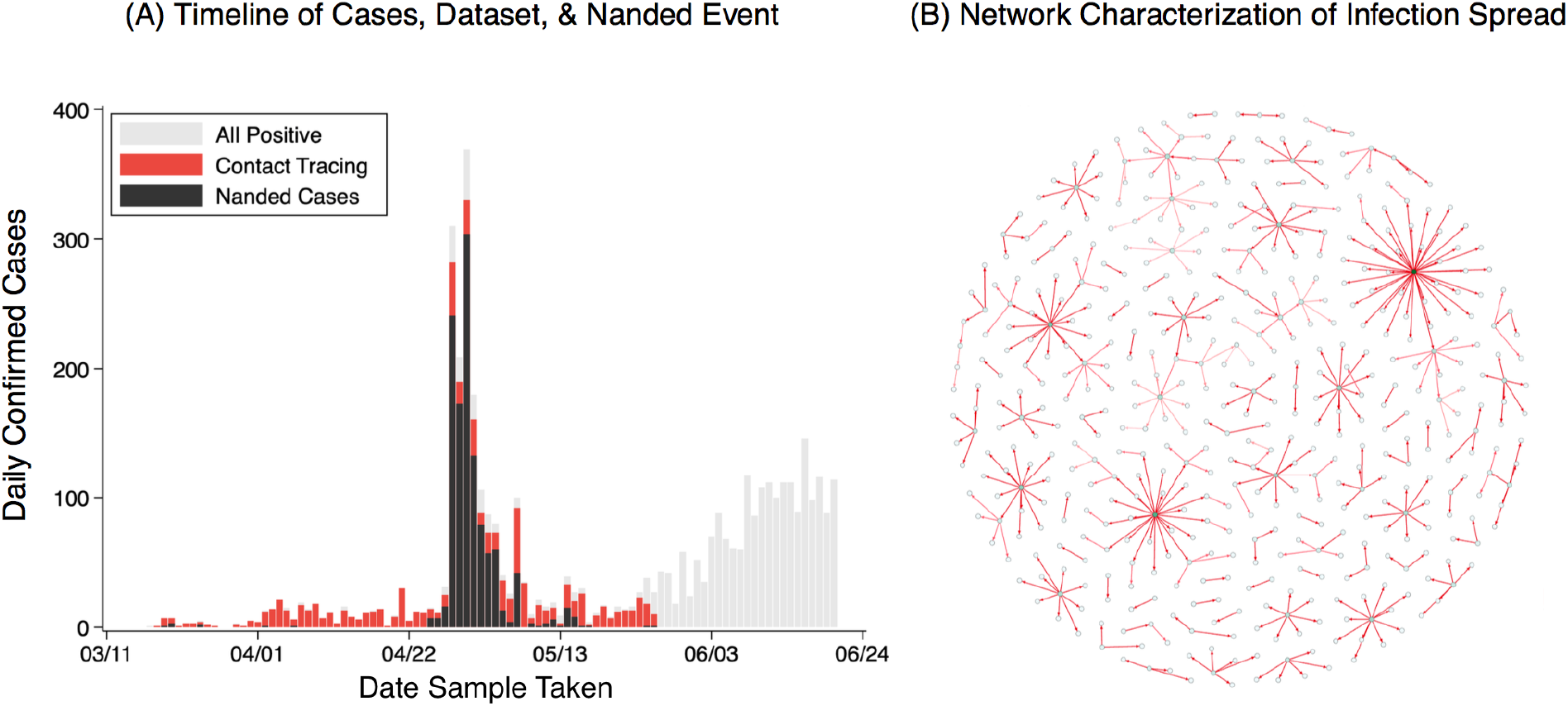
The data from Punjab. (A) Timeseries of reported cases in Punjab during the period of lockdown in the state (red bars) and those due to the Nanded event (black bars), and total cases from early March to the middle of June. (B) Visualisation of case clusters in the dataset, and their linkages from self-reported contacts. This network-type graph requires assumptions (see Materials and Methods). Most individuals infected only few others, while a few infected many: overall, 10% of cases accounted for 80% of infection events.

The data includes 454 initial cases and 11309 high risk contacts (Fig.1B). Confirmed cases comprise two groups: those residing in Punjab and who were likely infected within the state, and those who are thought to have acquired infection outside the state, due to travel or migration. Our analysis focuses on the former group, and in particular on *seeds* (the first infection in a cluster) in this group, these being the individuals amongst whom contacts are most clearly defined (see Materials and Methods). This yields a total of 148 seeds with 2763 contacts, although we also present sensitivity analysis when analysing all 454 seeds with at least one contact (and all 11309 contacts) in this data, a significant proportion (36%) of whom were religious pilgrims who returned to Punjab from Nanded, Maharashtra, after being stranded there for a month.

## Heterogeneity in transmission

The “secondary case distribution” is the distribution for the number of onward infections caused by an infected individual. We observe both the number of secondary cases for each individual, and the total number of contacts the person has. In mathematical modelling of transmission dynamics, heterogeneity in transmission is conventionally captured through modelling the secondary case distribution with a negative binomial distribution, allowing for extra-Poisson variation (*1, 5*). Fig. 2A illustrates the secondary case distribution in the data from Punjab. An important feature in this distribution, consistent with earlier findings (*4*), is that the majority (76%) of infected cases shows no evidence of onward transmission amongst any of their contacts. The negative binomial distribution captures these individuals, as well as the right-hand tail of the distribution, for example the 10% of individuals accounting for about 80% of transmission in this data. However, this distribution conceals further levels of heterogeneity, that can be important for epidemiological outcomes.

**Figure 2:**
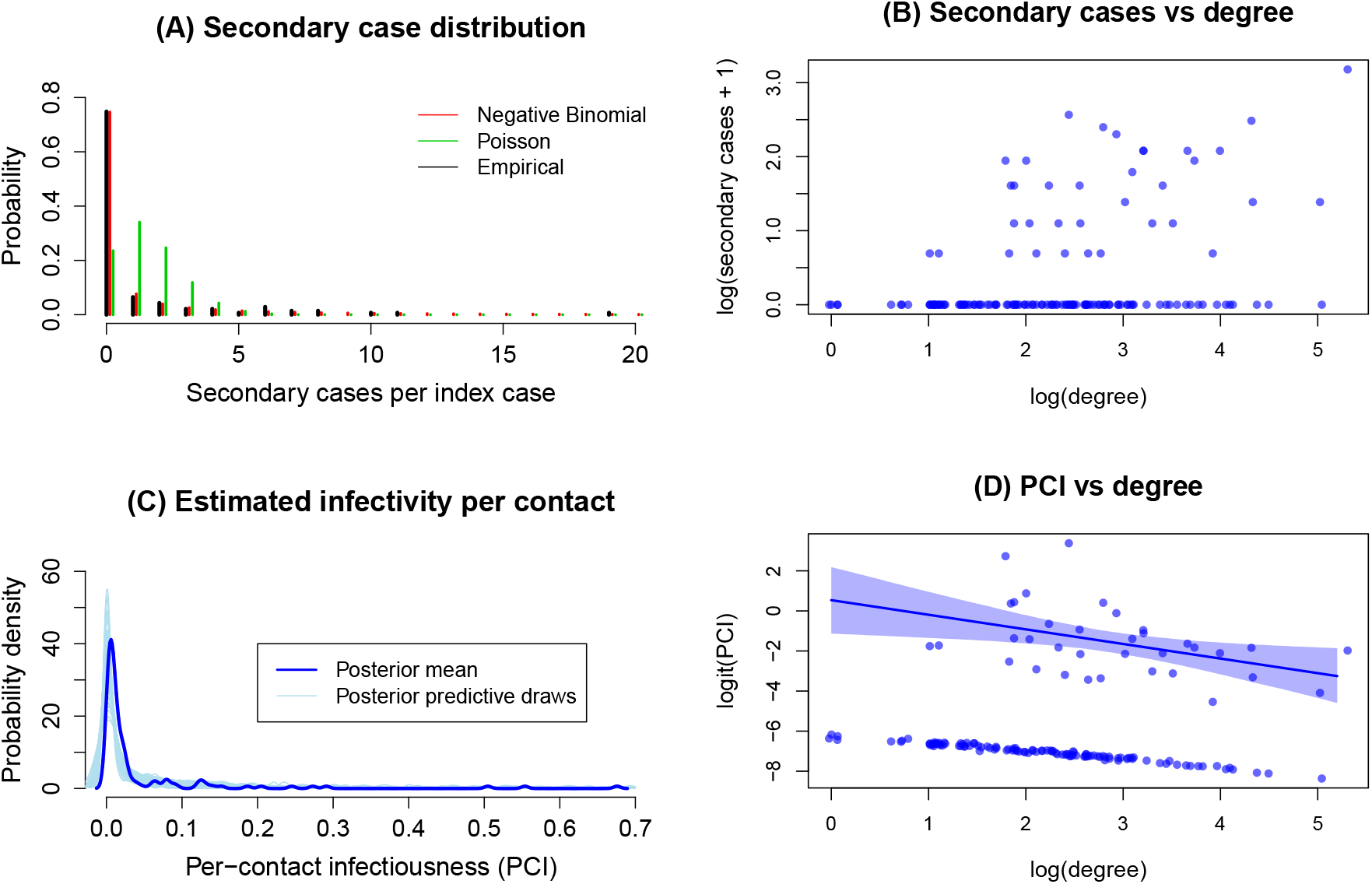
Heterogeneity of the data in secondary cases, and in numbers of contacts. (A) The distribution of secondary cases amongst ‘seeds’ (i.e. first cases in each cluster shown in Fig. 1B). Also shown, for comparison, are the best-fitting Poisson distribution (with λ = 1.4)), and the best-fitting negative binomial distribution (with distribution parameters *r =* 0.067, *k =* 0.1). The difference between the latter two curves illustrates the strong extra-Poisson variation in the secondary case distribution. (B) Scatter plot of secondary cases vs degree, at the individual level. The secondary case and degree distributions are shown at the logarithmic scale, and adjusted by 1 to account for zeros, to address skewness of the distributions. Although both secondary case and degree distributions show a strong right-skew (panel A), this figure illustrates that the latter does not explain the former: despite a positive relationship between the two distributions, a substantial number of individuals with low degree generate some infections, while many with high degree generate zero onward infections. (C) Estimated marginal density of per-contact-infectiousness (PCI) that, alongside degree, is needed to explain the heterogeneity in secondary cases. Shaded intervals show 95% Bayesian credibility intervals. (D) Estimated PCI vs degree. The figure displays relationship between the logarithm of the odds (logit) of PCI and the logarithm of the degree. These transformations allow us to plausibly model the joint distribution of PCI and degree as a multivariate normal in section 4 (see Materials and Methods and Supporting Information). There is a discernible lower band due to a large number of cases with zero onward infections, which have very low estimated PCI. Among those with onward infections, there is a discernible negative association.

Heterogeneity in transmission can arise from both biological and behavioural factors, including connectedness (the individuals with the most contacts having the most opportunities for transmission), and individual-level variation in infectiousness (for example, with between-individual and temporal variation in viral shedding (*9, 10*)). Fig. S2 (supporting information) illustrates the distribution in the number of reported contacts per infected case (the ‘degree distribution’) in our dataset, showing a pronounced right-skew similar to that of the secondary case distribution. However, this skew alone cannot explain the heterogeneity in the secondary case distribution: Fig.2B shows that there are many individuals in this data set who caused no further infections despite having many contacts (i.e. having ‘high degree’), and conversely many individuals with low- and moderate-degree who caused several onward infections. These data suggest that there is further heterogeneity acting at the individual level, modifying the effect of the degree distribution (see also Fig. S3).

To capture this heterogeneity we defined the ‘per-contact infectiousness’ (PCI) as the probability that a given contact results in infection, a probability assumed to vary by index case, but to apply equally to all contacts of a given index case. As shown in Fig.2B, there are several individuals with 1-2 contacts who caused zero onward infections, giving rise to substantial uncertainty in their true PCI (similar challenges apply to low-degree individuals who infected all their contacts). To address this issue we treated PCI as an individual-level effect and estimated it using Bayesian shrinkage, a technique employed (among other places) in the education statistics literature to estimate teacher effectiveness (*11-13*). Fig.2C shows resulting estimates for the marginal distribution of PCI over the population, once again illustrating a strong right-skew. Fig.2D illustrates this association between degree and PCI, showing: (i) a bimodal relationship between the two, arising from the large proportion of individuals that do not infect any others, and (ii) amongst those that do infect others, a negative association between degree and PCI. Overall, these findings illustrate that degree and PCI operate in tandem to drive heterogeneity in the secondary case distribution. Performing these analyses on the full data for seeds (including returnees as well as the ‘core’ group) shows qualitatively similar results (see Fig. S4). We next examined the implications of these associations, for transmission dynamics.

## Implications for transmission dynamics

We asked: (i) how important are the zero-infectors in these distributions, for epidemiological dynamics? (ii) How do outbreak dynamics compare when taking the conventional approach of using the secondary case distribution alone (Fig. 2A) vs when modelling both PCI and degree separately (Fig. 2D)? To address (i), we used the Poisson and negative binomial distributions shown in Fig.2A, the former being an example of capturing the mean secondary cases but failing to capture the proportion zero-infectors. To address (ii), we additionally modelled the log-transformed degree and logit-transformed PCI as following a bivariate normal distribution, with correlation *ρ* (see Table 1, and Materials and Methods). Consistent with Fig.2D, we assumed a range of values for *ρ*, from −0.4 to 0.

**Table 1:** List of the different models used, for capturing heterogeneity in the population. ‘Secondary case distributions’ (models 1 - 2) are as in Fig. 2A. They ignore any interactions between degree and PCI, and instead aim only to capture variation in the numbers of secondary cases per index case. By contrast, ‘Joint distributions’ aim to model the associations shown in Fig. 2D. They employ the bivariate normal distribution described in the Materials and Methods, with correlation *ρ*.

| Model number | Description |
| --- | --- |
| 1 | Secondary case distribution using Poisson distribution with mean 1.4 |
| 2 | Secondary case distribution using Negative Binomial distribution with number of successes = 0.1 and probability of success = 0.067 |
| 3 | Joint degree/PCI distribution with $\rho = -0.4$ |
| 4 | Joint degree/PCI distribution with $\rho = -0.2$ |
| 5 | Joint degree/PCI distribution with $\rho = 0$ |

For the transmission model we implemented a simple network simulation, in an assumed population of 3,000 individuals, consistent with the population size in this study. For simplicity and generality, we simulated the epidemic in generations of infection: our simulated outbreak behaviour would thus apply to any emerging infection sharing these heterogeneities (see Materials and Methods).

Fig. 3 shows a comparison of model projections for the behaviour of an index case: that is, when simulating only a first generation of infection. Results illustrate how it is possible to accommodate a wide range for *R_0_* (Fig. 3A), even amongst models that capture a high proportion non-infectors (Fig. 3B).

**Figure 3:**
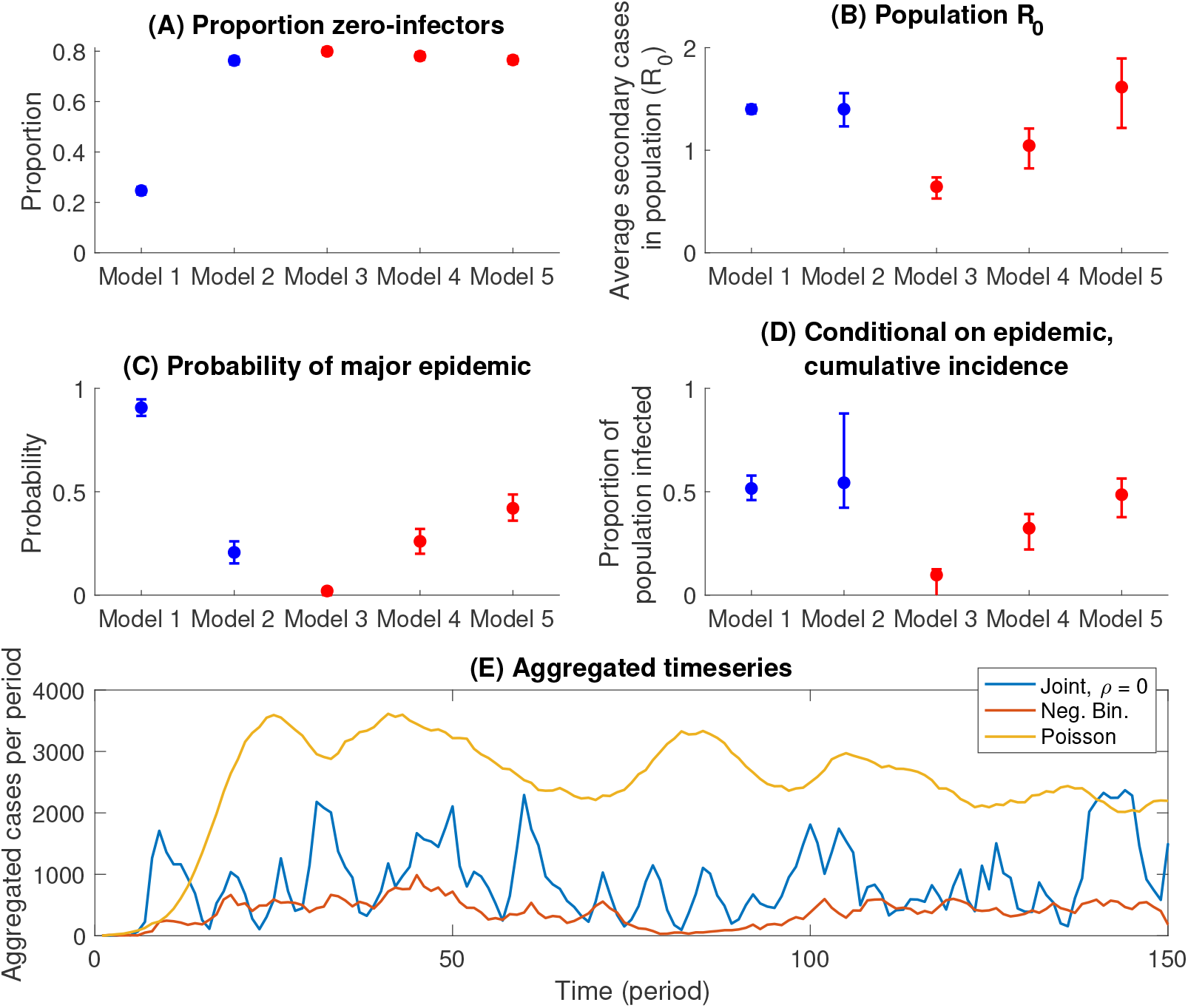
Results of simple transmission models incorporating heterogeneity. Top panels show the average behaviour of an index case in a fully susceptible population of 3,000: (A) The proportion of individuals that cause no further infections. (B) Distributions for the mean number of secondary cases caused by an index case, when averaged over the whole population. In each panel, blue points show outcomes when simulating only secondary case distributions, while red points show outcomes when simulating from the joint degree/PCI distribution described in the main text. Model numbers are as listed in Table 1. Of all models, only the negative binomial secondary case distribution, and the joint degree/PCI models capture the high proportion of index cases who do not cause secondary cases (panel A). However, even amongst these models, there can be substantial variation in *R*_0_ (panel B), owing to the role of correlation between degree and PCI. Middle panels (C,D) show epidemic outcomes over 500 time periods, assuming a 1% probability per time period, of exogenous introduction of an infectious case (here, an ‘epidemic’ is denoted as any simulation having a cumulative incidence > 500 cases (see Materials and Methods for rationale)). Uncertainty intervals arise from repeating simulations 250 times, and reflect 95% simulation intervals. (E) Modelled timecourse of incidence, when aggregated over 250 simulations (with each simulation being interpreted here as an independent location). A Poisson secondary case distribution (in yellow) gives rise to a large surge in aggregate infection because of epidemics in multiple locations occurring in a synchronised way.

Figs. 3C,D compare the outcomes of full epidemic simulations. By failing to capture the high proportion of zero-infectors (Fig. 3A), a Poisson secondary case distribution yields the most outbreak-prone populations, with 90% of simulations yielding major epidemics (Fig. 3C). Even amongst the remaining models, however, there is a notable disparity in epidemiological outcomes: amongst models capturing the joint distribution between degree and PCI (Models 3 - 5), it is not possible to identify a value of *ρ* that matches most closely to the negative binomial model for secondary cases (Model 2). While the latter appears intermediate to Models 3 and 4 in Fig. 2C, it is intermediate to Models 4 and 5 in Fig. 2D.

Figure 3E compares selected models in terms of the aggregate temporal pattern that they predict, when aggregated over multiple independent locations. Under epidemics simulated using a Poisson secondary case distribution, there is a surge of infection across several locations at once, a scenario that would place severe demands on health resources. By contrast in outbreaks driven by distributions capturing the high proportion zero-infectors, aggregate epidemic dynamics are more characterised by a series of asynchronous peaks in different locations, overall making for a lower peak demand on health services.

## Efficiency of contact tracing

Although contact tracing plays an important role in the SARS-CoV-2 response, in resource-constrained settings such as India, its demands on the healthcare system can make it difficult to sustain. Motivated by our findings, we propose reframing contact tracing with the goal of efficiently identifying individuals with high PCI. In our data overall, we estimate that if an individual caused at least one onward infection, there is a 79% probability that they caused at least two onward infections. We thus propose a sequential strategy where, for every index case, a ‘pilot’ subset of only s randomly selected contacts is first tested; the remainder of contacts are then followed up and tested, only if there is a positive in the pilot subset. Such a strategy could substantially reduce the overall contact tracing effort, while still effectively identifying high PCI individuals. Fig. 4 shows results of simulating such a strategy 1,000 times on the full dataset of 454 cases, for a range of values of *s*. The figure illustrates diminishing returns in the fraction of infections found, beyond a pilot subset size of 10 contacts (Fig. 4A). However, even with a pilot subset size of only 5 contacts, it is possible to identify 80% of infections (Fig. 4A), with <40% of the contact tracing effort that was expended in this data (Fig. 4B).

**Figure 4:**
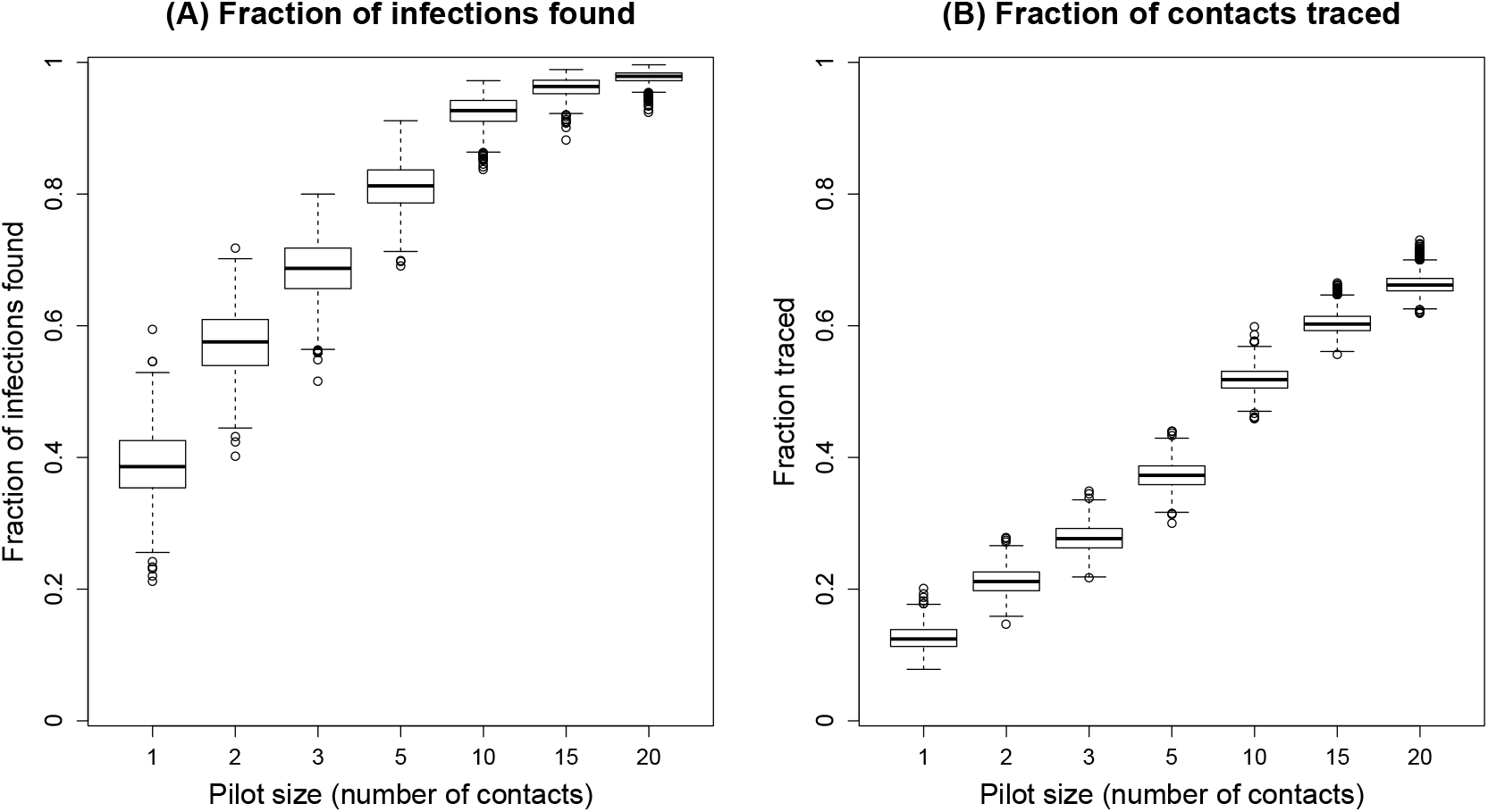
An approach to efficient contact tracing. Figure shows simulated outcomes of a strategy to test all contacts of an index case, only if there is at least one positive individual in an initial ‘pilot’ sample of *s* contacts. (A) The proportion of infections found as a function of s (B) Overall contact tracing effort, as measured by the proportion of contacts that would be traced, again as a function of s. Owing to the right-skew of the PCI, the left-hand panel illustrates diminishing returns with increasing s, suggesting, for example, that it would be possible to identify 80% of the cases in this dataset, with <40% of the contact tracing effort.

## Discussion

We have shown how individual-level data, gathered from the routine course of contact tracing, can be analysed to gain important insights into the transmission of SARS-CoV-2. As well as affirming findings from elsewhere, that the majority of cases appear not to infect any others (*1, 4*), our findings also highlight how heterogeneity in transmission may be more complex than previously recognised.

Simple dynamical models highlight the important role that is played by these heterogeneities. At the gross level of the secondary case distribution, the high proportion of zero-infectors yields outbreak dynamics wherein surges can be handled by providing mobile services rather than increasing hospital capacity in every geography (Fig. 3E). The negative binomial distribution, conventionally used in the modelling literature, captures this proportion well (Fig. 2A). However, our analysis also highlight some limitations of this distribution: accounting for underlying correlations between degree and PCI can lead to different outbreak dynamics, in terms of the risk and size of major outbreaks over time (Figs. 3C,D). Our results also have implications for the efficiency of contact tracing. When a large fraction of infected individuals do not cause onward transmission, we show the value of a simple two-step strategy, of, for example, first testing family members and then testing other contacts only if at least one family member is found to be positive (Fig. 4). Such approaches can be particularly valuable in resource-constrained settings such as India, in decreasing the requirements for contact tracing substantially, while still identifying most cases.

An important question, that we are not able to address using the current data, is what drives the heterogeneity in per-contact infectiousness. This heterogeneity may arise, for example, from biological factors such as the role of pre-existing, cross-reactive immunity that may moderate viral load in some individuals more effectively than others (*14*). Our analysis suggests that PCI increases with age and is significantly associated with sex (Fig. S5). Further data on these and other individual-level characteristics would be invaluable in further examining key risk factors for infectiousness. Where risk factors involve individual characteristics that can be readily identified in newly diagnosed patients, such as viral load, these factors could also play an important role in guiding future contact tracing efforts. However, heterogeneity in PCI could also reflect variations in the closeness of reported contacts, with some reporting only the closest contacts and others reporting wider contacts, thus explaining the negative correlation. We emphasise that our data is limited to defined ‘high-risk’ contacts (see Materials and Methods), thus excluding incidental contacts that might be expected to bias our estimates the most. Nonetheless, even if there is variation in closeness amongst high-risk contacts, our analysis offers an approach for adjusting for these variations, when interpreting what routine contact tracing data means for transmission: in this case our estimates for PCI should be regarded as a data-driven weighting of contacts, rather than infectiousness. Our approach can easily be adapted for any dataset where there is additional information on closeness of contact.

Amongst other limitations, the contact tracing data was collected, not under controlled study conditions, but as part of a public health response, by the Government of Punjab. Our approach to this data is pragmatic, recognising some inherent limitations: there may be false negatives in the data if people were tested too early, or indeed if people had been infected long in the past, which we cannot tell in the absence of serological tests. As with any contact tracing data, our assumptions for who-infected-whom, in a given contact pair, may be imperfect. We are able to address some of these concerns (for instance, by showing that our results are robust to a change in the directionality of a link [see Materials and Methods]). However, more-and better-data are absolutely necessary to refine our estimates, particularly on the nature of the correlation between degree and PCI. Further, although the lockdown conditions facilitate an indepth analysis of transmission amongst contacts, our findings must be interpreted with caution in scenarios with uninhibited transmission, as might occur in the absence of a lockdown or other non-pharmaceutical interventions (*15*). Additional limitations on the modelling are described in the Materials and Methods.

Overall, the methods that we have outlined here should apply to any contact tracing database and our publicly available code can be directly applied to any such data that have been collected. Contact tracing forms an integral part of the response to SARS-CoV-2 around the world: while being an important public health strategy in its own right, it can also provide invaluable information about how, and to whom, infection is being spread. Systematic analysis of this data could provide important insights to inform future, smarter strategies for the control of SARS-CoV-2.

## Data Availability

All data referred to in the manuscript are available on request from the authors

## Acknowledgments

NA was supported by the UK Medical Research Council and by the Bill and Melinda Gates Foundation. PM is grateful for support from a Ford Foundation grant, which supports work on use of tacit knowledge in urban environments. TM was supported by the National Institute of Mental Health of the U.S. National Institutes of Health under award number DP2MH122405. All authors contributed equally, and are listed in alphabetical order. The authors gratefully acknowledge Dr. Rajesh Bhaskar, who was responsible for the production of the dataset and generously shared it for analysis, Dr. Rajesh Bhatia, Dr. K K Talwar, advisors to Government of Punjab, Ms. Vini Mahajan, Ms. Isha Kalia, Ms. Tulika Avni Sinha, and Ms. Yamini Aiyar. The authors also acknowledge valuable support in collating and organising the data, from Vidisha Mehta, Kanhu Charan Pradhan, Harish Sai, Shamindra Nath Roy (Centre forPolicy Research), Olivier Telle (Centre for Policy Research/Centre National de la Recherche Francaise), and Benjamin Daniels (Georgetown University).

## Supplementary materials

Materials and Methods

Figs. S1 to S5

## Supporting information

### Materials and Methods

#### Contact tracing

The contact tracing, implemented by the Integrated Disease Surveillance Program in the state, was conducted in the following four steps (*16*).

1. Immediately after a confirmed case is identified, a trained epidemiologist or medical officer interviews the case and ascertains all contacts.
2. Contact tracing is then completed for all contacts who have interacted with the positive case anytime between 2 days prior to the onset of symptoms and the date of isolation, or a maximum of 14 days after symptom onset. So, if symptoms started on April 1st 2020 and the person was isolated on April 5th, all persons who were in contact with the case between March 30th and April 5th are to be traced. The data are listed with details of the contacts and this list is then shared with contact tracers for tracking.
3. The epidemiologist then classifies each contact as high- or low-risk. The definition of high-risk is those who face-to-face conversations for at least 15 minutes with the positive case or physical contact. Contacts who are out-of-state are passed on to other states.
4. High-risk contacts are then tested by a lab technician. Contacts who are negative and remain asymptomatic for 28 days are released from the list. For those who are positive, the listing is again initiated to trace a further generation of contacts.

In the data, we recoded the reasons for testing into six categories: Seed (Normal), Contact (Normal), Farmer/Labour, Migrant/Returnee (Non-Nanded), Nanded, Other. During the lock-down, there was a fear that those entering from elsewhere would bring the infection to Punjab. Pilgrims returning from Nanded (described in section 2), other migrants/returneees, and farmers/laborers (residing in Punjab but originally from outside it) were tested by a special protocol. The “other” category consisted of certain high-risk populations like frontline healthworkers who were tested for occupational reasons and their families. The categories “Seed (Normal)” and “Contact (Normal)” correspond to those tested due to normal protocol - usually due to symptoms, living in a containment zone, or from the contact-tracing protocol described above - as the first case in a cluster or the contact of a confirmed case, respectively.

Table 2 displays the frequency of each reason and the percentage reporting no high risk contacts, with distributions of gender, age and symptomatic status. As we might expect laborers and migrants tend to be younger and more male. Of particular concern is the high percentage of individuals reporting no high risk contacts in most categories (Zero Deg). This is due to a bookkeeping problem. For contacts of a previously confirmed case we can only retrieve the number of high risks contacts that had yet to be tested — so those contacts shared with the person from whom the infection was contracted are not counted. And returnees from elsewhere often have no contacts listed in Punjab.

**Table 2:**
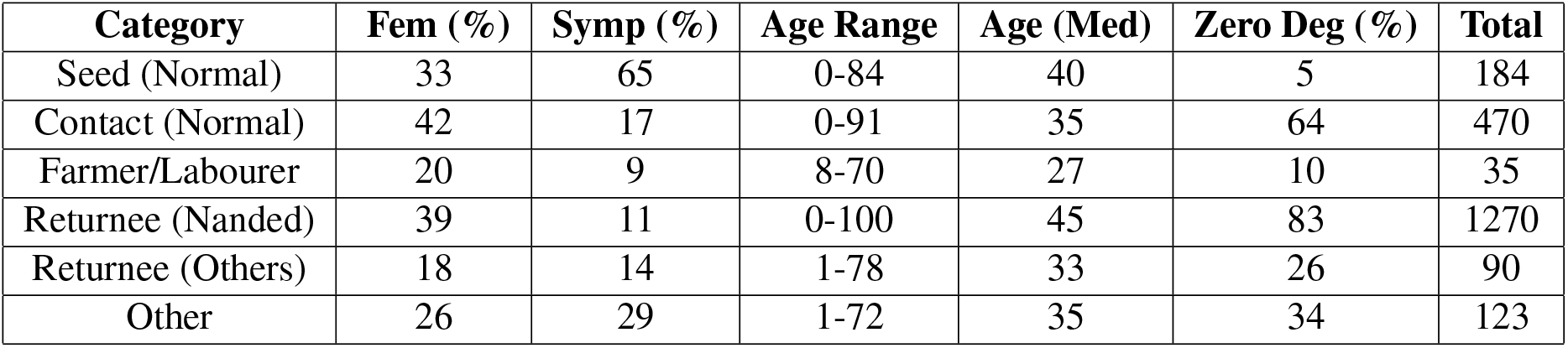
Key Sample Descriptives. We provide sample descriptives for six reasons for testing: Seed (Normal), Contact (Normal), Farmer/Labour, Migrant/Returnee (Nanded), Migrant/Returnee (Others)[those not in the Nanded event], Other. We provide information on the percentage of people in each category who are female (Fem), show symptoms (Symp), and report no high risk contacts (Zero Deg). We also display median age (Age (Med)) and the minimum to maximum age (Age Range) for each category. Missing observations are removed in this table.

| Category | Fem (%) | Symp (%) | Age Range | Age (Med) | Zero Deg (%) | Total |
| --- | --- | --- | --- | --- | --- | --- |
| Seed (Normal) | 33 | 65 | 0-84 | 40 | 5 | 184 |
| Contact (Normal) | 42 | 17 | 0-91 | 35 | 64 | 470 |
| Farmer/Labourer | 20 | 9 | 8-70 | 27 | 10 | 35 |
| Returnee (Nanded) | 39 | 11 | 0-100 | 45 | 83 | 1270 |
| Returnee (Others) | 18 | 14 | 1-78 | 33 | 26 | 90 |
| Other | 26 | 29 | 1-72 | 35 | 34 | 123 |

Accordingly, in the main text we restrict our analyses in the text to the 148 seeds tested during normal protocol that do not have a missing of zero value in the number of high risk contacts, as contacts are required to estimate PCI. Naturally, because they have come through the normal protocol, seeds have a much higher proportion of symptomatic individuals (Symp). This is a population for whom we believe we have a robust contact distribution applicable to the population of Punjab and for whom we can reliably identify seeds and contacts.

Where such information could be ascertained, we undertook an extensive exercise of matching contacts to seeds in the entire dataset, to verify the dataset had been coded correctly. Nonetheless, we were concerned about the *case ascertainment* problem. Assume that both A and B have tested positive. *B* could have infected *A* (directly or indirectly) but we observed *A* first and coded it as a seed. Two possibilities exist either we got it right and A is the seed, or we got it wrong and *B* is the seed. While we can never be sure of this answer, we can test the robustness of our claims to swapping seeds and contacts. While we cannot direct compare onward infections and degree of contacts to seeds due to the bookkeeping problem, we can test whether seeds and contacts display similar infectiousness. Indeed, we see that that the contacts (2763 individuals) of normal seeds have an aggregate test positivity of 6.0% while the contacts (1885 individuals) of normal “contacts” have a test positivity of 6.5%. These are statistically indistinguishable (*p =* 0.45). Thus, as long as the coding of seed and contact by the government of Punjab was independent of degree (which is likely because seeds were typically tested due to a biological criterion - showing symptoms - and not a social criterion), we surmise that our estimates of the secondary case distribution are likely to be robust to swapping seeds and contacts.

Beyond the core dataset, as table 2 shows, the majority of positive cases are from the Nanded event, from which pilgrims were brought back on dedicated buses. This group is akin to the Diamond Princess experience, where multiple people were in contact with each other in close quarters. As such, seeds are contacts were not well-defined in this population.

Nevertheless, in Fig S4 in the supporting information, we also show robustness when extending our analysis to the 454 seeds across the Nanded event and all other categories (as seeds are also present in each of the four special protocol categories) who report at least one contact.

#### Bayesian shrinkage

A natural estimate of PCI for person *i, p_i_* would be to divide the number of onward infections (*z_i_*) by the number of contacts (*d_i_*), i.e., 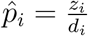. The shape of the degree distribution presents a challenge for this method, however, as it means the variability in the estimated PCI varies across individuals based on their number of contacts. As an example, consider two individuals, *A* and *B*, with 2 and 100 contacts, respectively who have infected no one. We are confident that *B* has a PCI close to zero but not so confident with A due to a small sample size. We address this issue through *Bayesian shrinkage*. In this setting, individual estimates of PCI (*p_i_*) from high contact individuals (such as *B*) will be mostly unchanged while those from lower contact individuals (such as A) will be shrunken towards the overall mean (*17*).

Amongst different ways of performing Bayesian shrinkage (e.g., the Beta-Binomial model), we chose to model the logarithm of the odds (logit) of the PCI as following a normal distribution with a common mean and variance, as this functional form is closely linked to our modelling of transmission dynamics (other approaches yield similar estimates (*18*)). In particular, we estimate:

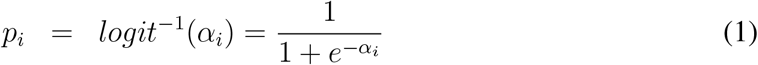

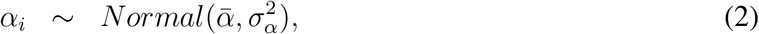

where *α_i_* is the log-odds of *p_i_* and 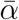 is the common mean. Above, 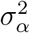 is inversely correlated to the amount of shrinkage. As 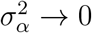, each *α_i_* is given the same value, so each *p_i_* is estimated as the mean infection rate. As 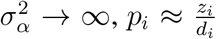. In practice, the “hyperparameters” like 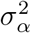 and *α* are estimated using Markov Chain Monte Carlo (MCMC) methods with diffuse priors. The non-zero values of 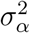 and 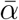 guarantee that our estimated *p_i_* is between 0 and 1.

In future work with additional data, it may be possible to characterize the complete joint distribution of PCI and number of contacts, thus alleviating the need to shrink to a common mean. For example, healthcare workers with training in mitigating infectious disease spread may have many contacts, but lower PCI than would be expected from the rest of the population. In such a setting, shrinking towards a single mean would underestimate the heterogeneity in the PCI distribution. Most of the extreme cases are 0’s in the data from Punjab, however, so in practice these values will be shrunk towards a small, non-zero value under either model.

#### Mathematical modelling of transmission dynamics

We implemented a simple network simulation, in an assumed population of 3,000 individuals (consistent with the population size in this study). For simplicity we modelled all networks as random, that is, neglecting clustering and other forms of network structure of higher order than the degree distribution. Also for simplicity, we simulated the epidemic in terms of generations of infection, rather than in continuous time: our projections could be interpreted as being conducted in discrete time, with a time interval corresponding to the mean generation time. The focus of this modelling analysis is to understand the importance of degree distribution and PCI for transmission dynamics in general; we thus did not model the details of symptomatic vs asymptomatic infection for SARS-CoV-2, nor of the pronounced variation of severity by age (*19*).

##### Network construction

For the Poisson and negative binomial secondary case distributions in Table 1, we drew 3,000 samples. We then constructed a random, directed network treating these samples as degrees, to construct a network of the secondary cases that any given individual would cause, once themselves infected (our results in Fig. 3 are qualitatively unchanged when assuming a directed network instead).

In figure S2, we show that the degree distribution in the data follows an approximately log normal distribution, and in our discussion of Bayesian shrinkage (above) we showed that the logarithm of the odd (logit) of PCI is constructed to follow a normal distribution. We note further that Fig. 2D and Fig. S4(D) show that the correlation between the log-transformed degree (*n*) and logit-transformed PCI (*p*) is plausibly negative for those that infect others. We consider a population of infected individuals. Since the log-transformed degree and logit-transformed PCI each follow a normal distribution and may be correlated, the natural choice for the joint degree/PCI distribution is to model the log-transformed degree and logit-transformed PCI as following a bivariate normal distribution:

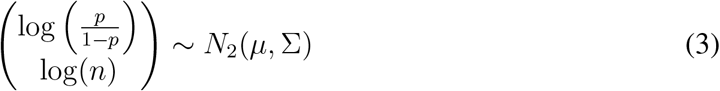

where *μ* is a vector composed of the mean values for logit-transformed PCI and log-transformed degree, and we have, for the covariance matrix ∑:

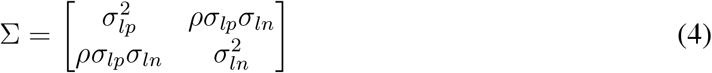

where *σ_lp_* is the standard deviation for the logit-transformed PCI; *σ_ln_* is the standard deviation for the log-transformed degree; and *ρ* is the correlation between the two. This construction allows us to explore different hypothetical scenarios for the correlation, and their implications for outbreak dynamics, while maintaining the correct shapes of the marginal distributions for degree and PCI. We posed three scenarios for *ρ*, taking values of −0.4, −0.2 and 0. The model we use for simulation differs from our Bayesian shrinkage approach presented in Fig. 2D. This distinction is necessary because, in our simulations, we wish to explore variation across multiple similar contact and PCI distributions. In any data analysis, however, we will model *conditional* on a particular observed contact distribution.

In a given simulation, we then sampled 3,000 values for degree and PCI. We constructed a random, undirected network from the degree distribution. For each individual m, we assumed that the sampled PCI *p*(*m*) applies uniformly to all of their contacts. Thus, although the link between any two individuals A and B is undirected - representing a bidirectional transmission risk - the transmission intensity is not necessarily the same in both directions, and depends on the respective PCIs of A and B (owing to between-individual variations in infectivity).

##### Epidemic simulation

For a given population constructed as above, suppose *C_t_* is the set of individuals that are infective at the beginning of time-step *t; S_t_* is the set of individuals that have not yet had infection and are therefore susceptible; and *J_t_* is the set of individuals that are newly infected in timestep *t*. Further, suppose that *p*(*m*) is the sampled PCI for individual m. Then we proceeded along the following iterative steps:

<u>While *t ≤* 500 and *S_t_, C_t_* both have at least one member:</u>

1. Identify *C_t_* with *J_t_*_−1_, and initialise *J_t_* as an empty set
2. For every member *m* of *C_t_*:

- Determine all contacts of *m* who belong to *S_t_* (for Models 1,2, regarding ‘contacts’ as secondary cases).
- For each such contact, conduct a Bernoulli trial with probability *p*(*m*), to determine whether infection occurs (for Models 1,2, taking *p*(*m*) = 1).
- Accumulate all new infections thus occurring in *J_t_*, and remove them from *S_t_*.
3. Perform a Bernoulli trial on all members of *S_t_* with probability 0.01, to identify exogenous introductions of infection. Accumulate all new infections thus occurring in *J_t_*, and remove them from *S_t_*.
4. Increment *t* by 1, and iterate from (1).

**Figure S1:**
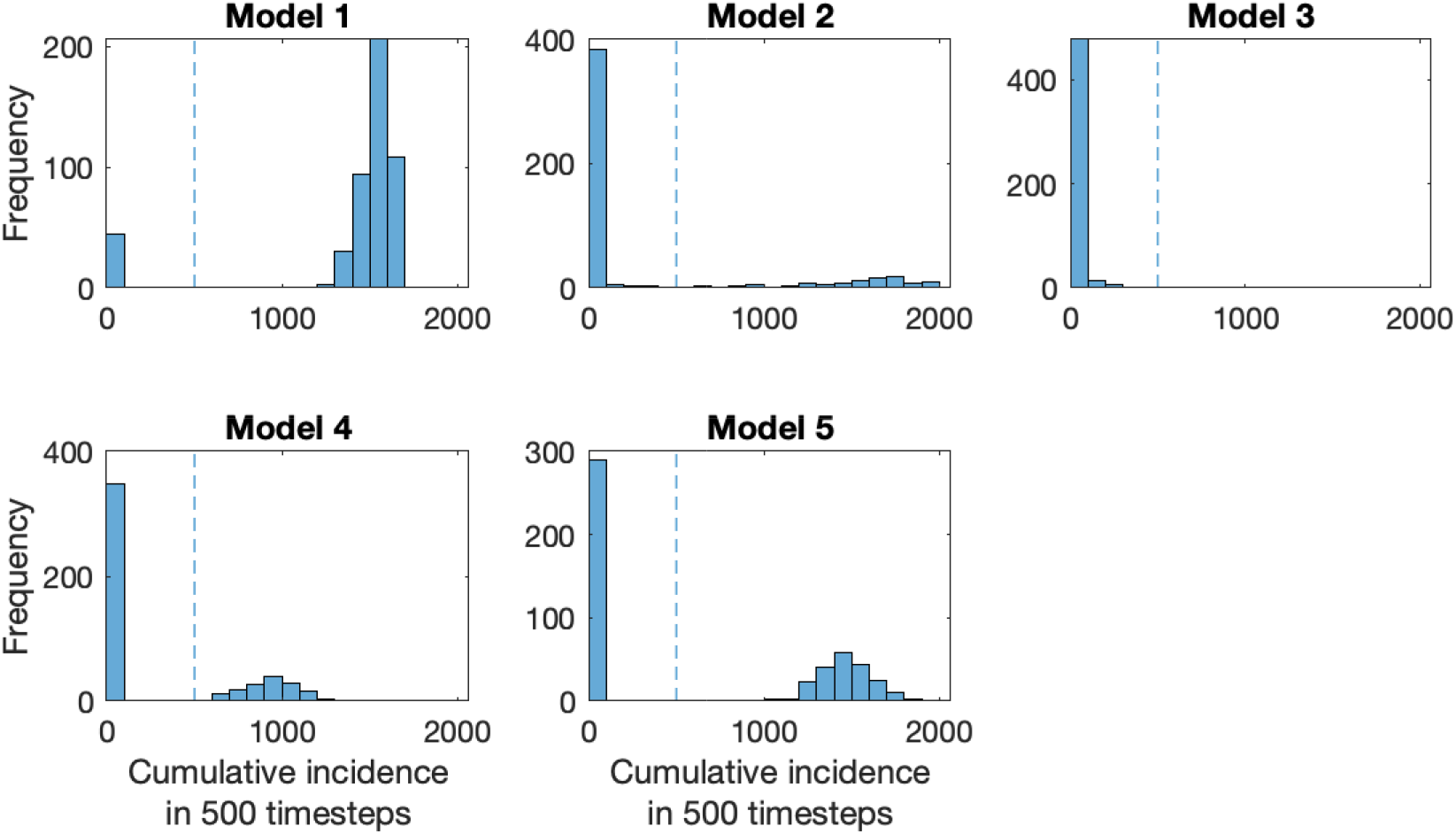
Frequency distributions for cumulative incidence, over 500 timesteps, for each of the models listed in Table 1 in the main text. Vertical dashed lines indicate a cumulative incidence of 500, a consistent dividing line between the two modes in these distributions. Thus Fig. 3C in the main text shows the probability mass to the left of this line, while Fig. 3D shows the mean cumulative incidence to the right of this line.

We repeated this algorithm 250 times, for each of the models listed in Table 1. Figure S1 shows the frequency distributions for the cumulative incidence thus obtained, i.e. the total cases over all 500 timesteps. The figure illustrates a bimodal pattern of outbreak sizes, with the vertical dashed line (at 500 cumulative cases) illustrating a consistent dividing line between the two modes. Accordingly in Figs. 3C,D in the main text, we denote ‘major epidemics’ as any simulation in which cumulative incidence exceeds 500 cases.

##### Limitations

For simplicity we have adopted a simple network model that models the progression of an epidemic through generations of infection. This simplicity is helpful for focusing the model-based analysis on the specific types of heterogeneity revealed by our analysis of the data. It also has the benefit of generality, showing epidemiological behaviour that would apply to any disease with the same underlying heterogeneity, regardless of the details of natural history. However, an important area for future modelling work would be to incorporate some important characteristics in the natural history of SARS-CoV-2, such as symptom status, age, and the full spectrum of severity of infection (*19,20*). Such refined models would be important, for example, in translating our simulated dynamics to timescales more specific to SARS-CoV-2. As mentioned above, we also take a simplified approach to the network structure, assuming the simplest case of a random network, and thus ignoring the potential for clustering, or other types of network topology that could be influential in transmission dynamics (*21*). Further data on the underlying contact structure, including the retention of information on test-negative contacts, would be helpful in addressing these simplifications.

#### Modelling efficient contact tracing algorithms

We used the following algorithm to perform the simulations for Figure 4. First, we choose s, the number of ‘pilot’ contacts to be tested. Then, for each of the cases in the data from Punjab take the number of observed cases and contacts and create a vector of 1’s and 0’s to designate, respectively, infected contacts and those not infected. We randomly assign the position of each of the infected cases in the vector and set the number of infected cases equal to the number observed in the data for that case. We then take s samples without replacement and with uniform probability from the constructed vectors. If at least one of the sampled units is positive, we simulate testing the remainder of the contacts (meaning that the number of tests equals the observed number of contacts and the number of infections found equals the observed number of infections). If none of the s sampled contacts are positive we do not do further sampling, meaning the number of infections identified is zero and the number of tests is *s*. We repeat this exercise across all cases in the data from Punjab 1000 times for each value of *s*.

Our simulation is illustrative, with some caveats to note. First, the procedure is not optimized for the number of contacts to test. The problem we address is similar to a bandit problem in the machine learning literature where, as more information about the PCI distribution is available through testing, the number of contacts to test is optimized to maximize the (expected) number of infections found while minimizing the number of tests. We anticipate that this sequential procedure would be challenging to implement in a public health context, particularly in a low resource setting. Instead, we opt for a simple rule that can be implemented with no additional optimization (e.g. testing family members and only testing further if at least one is positive) that can still substantially improve efficiency. We also need information about the joint distribution of PCI and contacts for a fully optimized approach, which we cannot estimate with precision in the data from Punjab. Additionally, we do not consider imperfect tests. False positives would preserve the number of infections found but make the procedure less efficient since some individuals will be tested based on false positives that would not otherwise be tested. False negatives will reduce the number of infections found, though this impact would be mitigated by the right-skew of the PCI distribution. Finally, this procedure relies on the availability of tests with relatively rapid results and the willingness of individuals to be tested. If the delay between testing and receiving results is too long, then contacts who were not tested in the pilot stage could be infecting others in the population while waiting to be tested.

## Supplementary Figures

**Figure S2:**
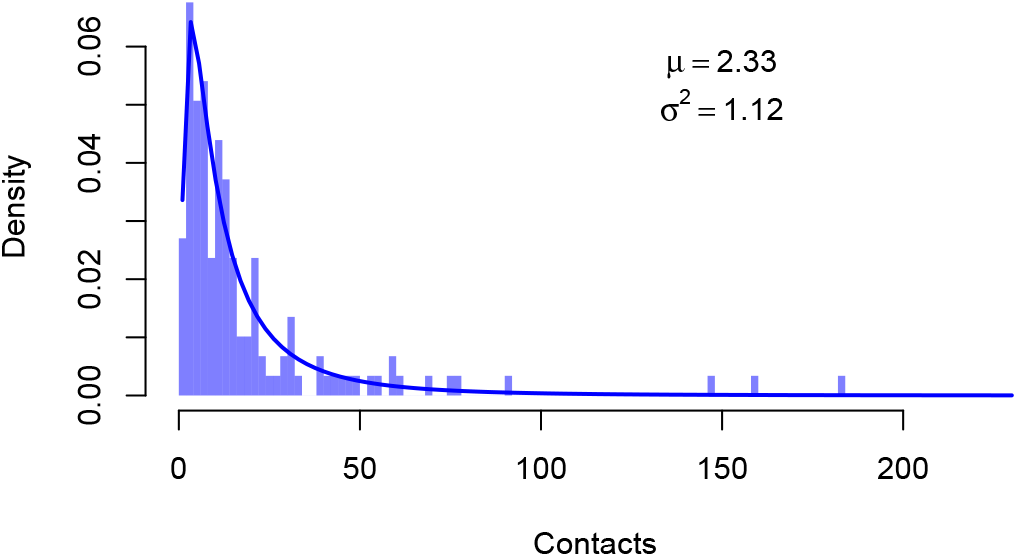
The Contact Distribution. The light blue bars denote the histogram of contact distribution with a bin size of 2. The density function of the log normal distribution with *μ* = 2.33 and *σ*^2^ *=* 1.12 fits the empirical distribution well. Consistent with standard network structure, this distribution has a strong right skew.

**Figure S3:**
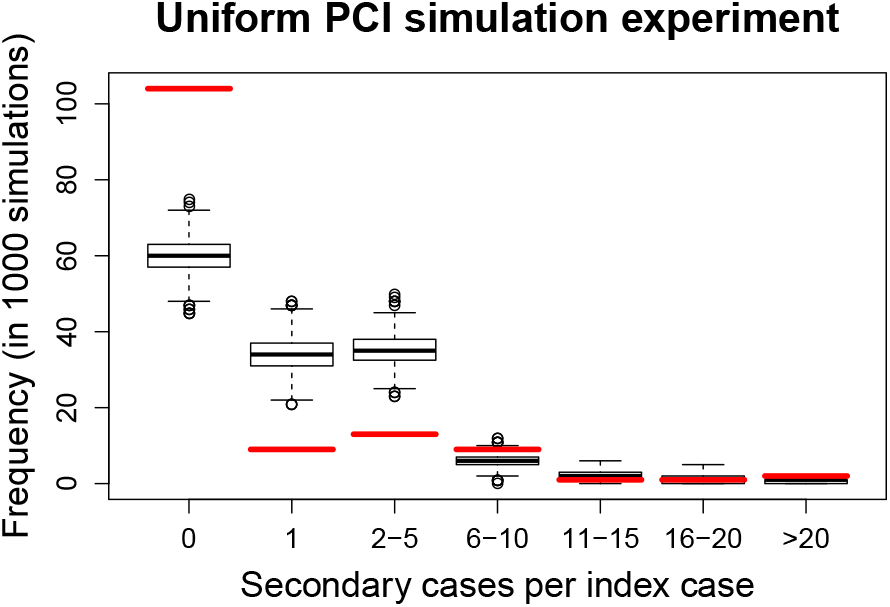
Inadequacy of the degree distribution to explain the secondary case distribution. While Fig. 2B in the main text illustrates this point visually, this plot offers statistical support. Red lines show the data for the secondary case distribution, while box plots show the bestfitting projections when assuming the degree distribution illustrated in Fig. S2, and moreover that the risk-of-transmission is constant across contacts (that is, a constant PCI). Doing so yields a secondary case distribution that severely underestimates the proportion of cases that caused zero onward infections.

**Figure S4:**
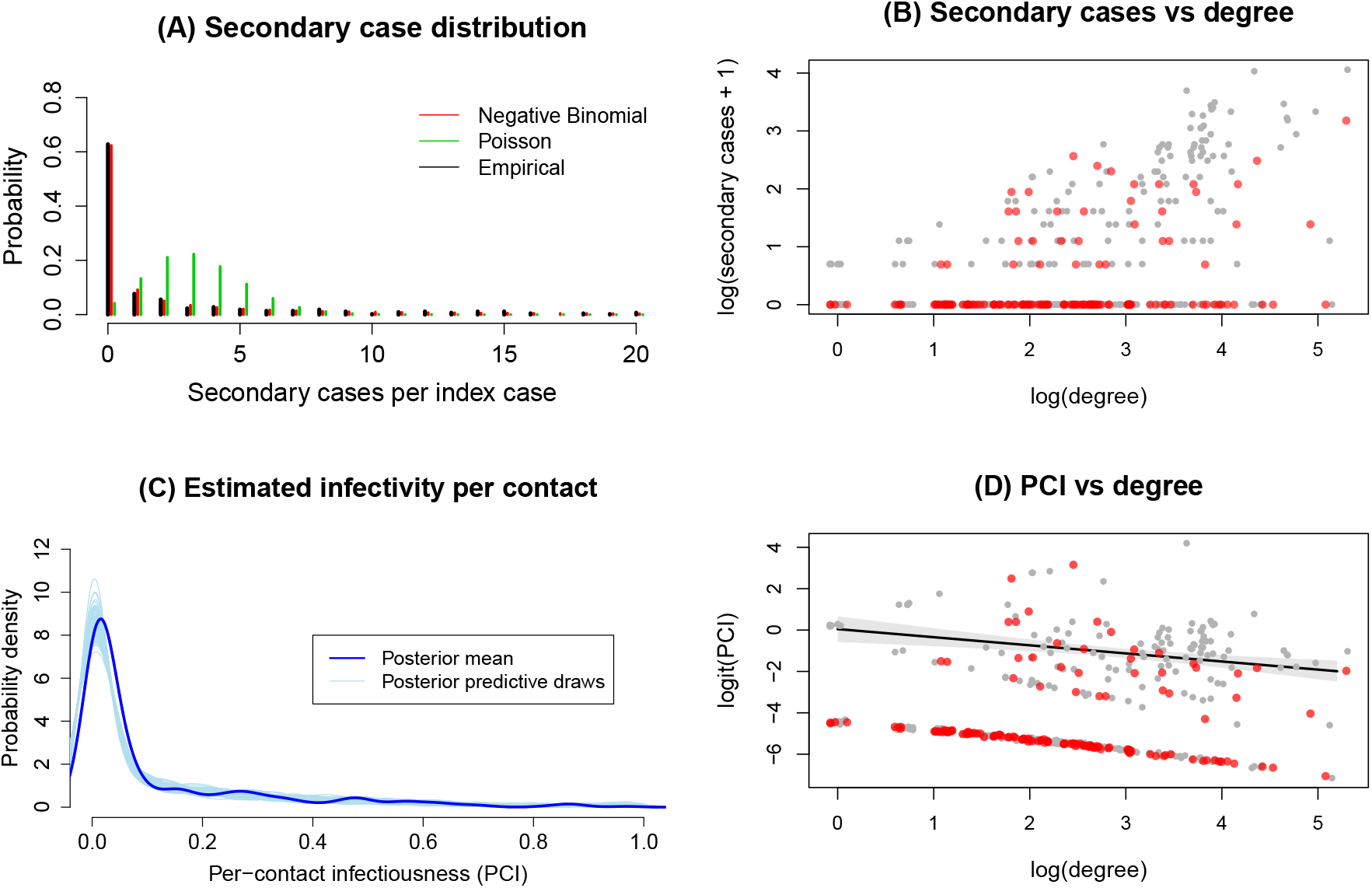
Robustness for full sample. The full heterogeneous sample is characterized in Materials and Methods. While the main text displays analysis on the core sample, the trends hold on the full dataset. Fig. S4(A) replicates Fig. 2A for the full sample, modelling negative binomial and Poisson fits to the secondary case distribution. As before, the negative binomial fits the secondary case distribution well, while the Poisson distribution underestimates the number of zero infectors. Fig. S4(B) shows a scatterplot of the logarithm of the degree distribution against the logarithm of the secondary case distribution adjusted by 1 to account for zeros and the skews in the distribution. The red points denote the core sample and the gray points denote the remainder of the sample. In both samples, although there is a discernible positive association between onward infections and degree, the heterogeneity in degree does not fully explain that of the secondary case distribution. Fig. S4(C) replicates Fig. 2C for the whole sample, showing the marginal distribution of PCIs. It is right-skewed as in the core sample. Fig. S4(D) shows the association between the log odds (logit) of the PCI and the logarithm of the degree. The red points denote the core sample and the gray points denote the remainder of the sample. Both samples are bimodal with a discernible negative association for those that infect others.

**Figure S5:**
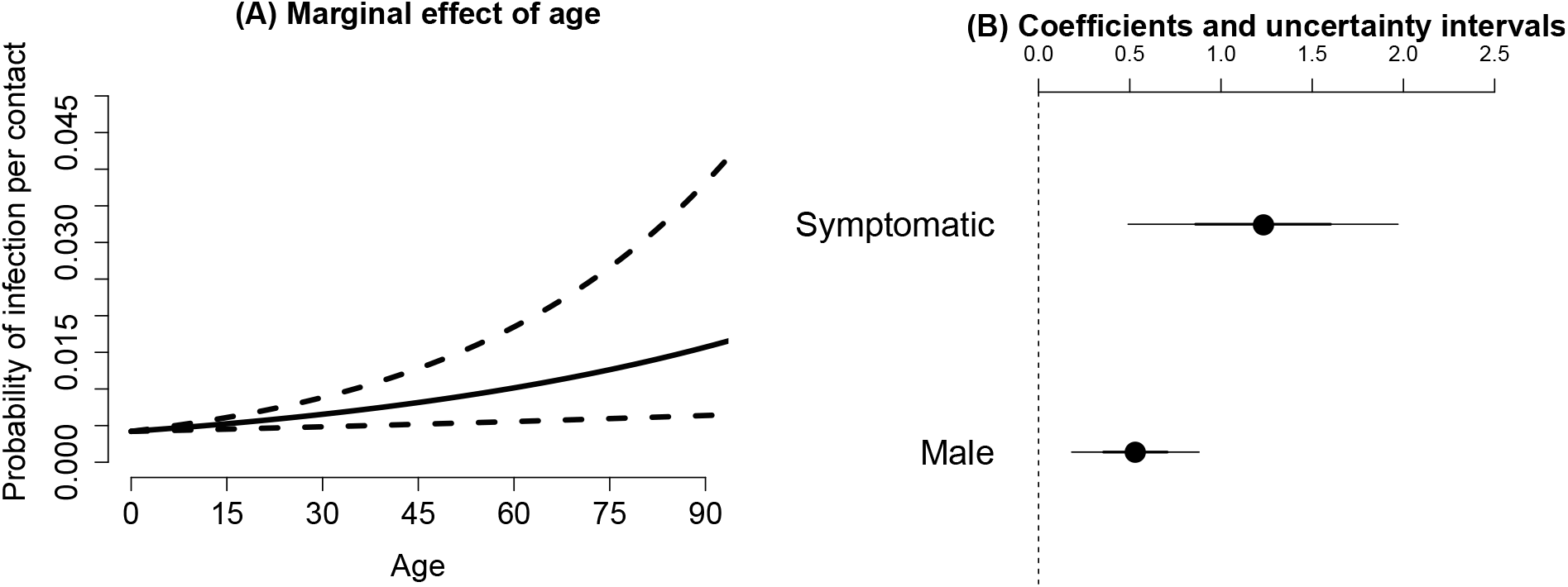
Regression results for likelihood of a secondary infection, against the age, sex and symptom status of the index case. Results are from a multivariate logistic regression. Panel (a) shows the marginal effect of age on the probability of secondary infection per contact for the reference groups (asymptomatic, female). Dashed lines represent the bounds of the 95% confidence interval on the coefficients. Panel (b) shows the logistic regression coefficients for the dichotomous variables associated with being symptomatic and being male. For a person at the average age (about 42 years old) being male raises the probability of secondary infection per contact by about 0.5 percentage points (from 0.8% to 1.3%). For a person at the average age being symptomatic raises the probability of secondary infection per contact by about 1.9 percentage points (from 0.8% to about 2.7%). Recall that these estimates come from data where the majority of individuals have no secondary infections, which reduces these overall probabilities.

## References

1. A. Endo, Centre for the Mathematical Modelling of Infectious Diseases COVID-19 Working Group, S. Abbott, A. J. Kucharski, S. Funk, Wellcome Open Research 5, 67 (2020).

2. Y. Wang, P. Teunis, Frontiers in Medicine 7, 329 (2020).

3. M. G. M. Gomes, et al., MedRxiv p. 2020.04.27.20081893 (2020).

4. R. Laxminarayan, et al., medRxiv (2020).

5. J. O. Lloyd-Smith, S. J. Schreiber, P. E. Kopp, W. M. Getz, Nature 438, 355 (2005).

6. T. Britton, F. Ball, P. Trapman, Science 369, 846 (2020).

7. J. Wallinga, M. van Boven, M. Lipsitch, Proceedings of the National Academy of Sciences 107, 923 (2010).

8. P. G. T. Walker, et al., Science 369, 413 (2020).

9. Y. Fu, et al., European Respiratory Journal (2020).

10. L. Qi, et al., International Journal of Infectious Diseases 96, 531 (2020).

11. R. Mendro, et al., Annual Meeting of the American Educational Research Association, San Diego, CA (1998).

12. J. Lockwood, et al., Journal of Educational Measurement 44, 47 (2007).

13. J. Lockwood, D. F. McCaffrey, L. T. Mariano, C. Setodji, Journal of Educational and Behavioral Statistics 32, 125 (2007).

14. N. Le Bert, et al., Nature (2020).

15. Report 9: Impact of non-pharmaceutical interventions (npis.

16. Ministry of Health & Family Welfare, India, Guidance document for POEs, states and UTs for surveillance of 2019-nCoV, Tech. rep. (2020).

17. A. Gelman, et al., Bayesian Data Analysis, Third Edition, Chapman & Hall/CRC Texts in Statistical Science (Taylor & Francis, 2013).

18. A. Gelman, J. Hill, Data Analysis Using Regression and Multilevel/Hierarchical Models (Cambridge University Press, 2006).

19. R. Verity, et al., The Lancet Infectious Diseases (2020).

20. Y. Liu, et al., The European respiratory journal 55, 2001112 (2020).

21. L. Danon, et al., Interdisciplinary perspectives on infectious diseases 2011, 284909 (2011).

